# The prevalence and management strategies of gestational urinary tract infections (UTI) in Kisumu County, Kenya

**DOI:** 10.1101/2022.06.17.22276561

**Authors:** Eunice Namuyenga Toko, Shivanthi Samarasinghe, Esther Furaha, Tariq Kapasi, Bertha Ochieng’, Collins Ouma

## Abstract

**Background:** Urinary tract infections (UTI) contribute to substantive proportions of adverse pregnancy outcomes. Current national statistics in Kenya show high maternal mortality (488/100,000) and neonatal mortality (24/1,000) rates. Kenya continues to report increasing prevalence and incidence rates of UTI associated with maternal and neonatal deaths. Kisumu County in western Kenya has a high maternal mortality rate of 495/100,000 with uncaptured maternal morbidity relative to the national average. However, information on the epidemiology of gestational UTI in the County, is limited. Semi-urban Chulaimbo and Nyahera Sub-County hospitals were used as model facilities to establish the burden of UTI during pregnancy and the specific clinical diagnosis and therapeutic management strategies.

**Methods:** Socio-demographic, laboratory and clinical history data was extracted from 416 pregnant women’s maternal child health data sets from health records between February 2019 to February 2020 using pre-designed data collection forms. Descriptive analysis was used to summarize the study population’s demographic characteristics. Chi-square test was used to establish proportionality. Qualitative data were thematically summarized. For all analyses, *P*≤0.05 was considered statistically significant.

**Results:** The study population had a mean of two (2) (±1.14) ante-natal (ANC) visits; a mean mothers age of 23.92 (± 6) years old; a mean parity of 2 (±2) and a mean haemoglobin level of 10.73(±1.8). About 56% (233/416) of the mothers attended the first ANC visit at varied gestational age. Only 1.4% (6/416) had a clinical history capturing UTI infection out of the total prevalence of 57.9% (241/416) diagnosed UTI positive by routine ANC profile deep stick urinalysis test. These clinical history data sets 1.4% (6/416) revealed a broad-spectrum therapeutic management of gestational bacterial infections using first line antibiotics.

**Conclusions:** Most UTI positive cases go without specific clinical diagnosis and management, posing a high risk of antibiotic drug resistance and development of specific bacterial allied gestational complications.

## BACKGROUND

Urinary tract infection (UTI) is the most common bacterial infection during pregnancy and can manifest as symptomatic or asymptomatic causing high morbidity and mortality rates in low-and-middle-income countries (LMIC) (2,3). Uncomplicated UTI present with frequency, urgency, dysuria, or supra pubic pain in women with a normal genitourinary tract (4). Complicated UTI is associated with functional or structural abnormalities of the genitourinary tract, with the involvement of either the bladder or the kidneys (4–8). Asymptomatic UTI is a persistent, actively multiplying bacterial infection within the urinary tract without any symptoms of infection. Its prevalence depends on parity, race and socio-economic status. Untreated asymptomatic bacteria may lead to development of acute symptoms of infections during pregnancy with serious consequences in form of fetal and maternal morbidity. For example, the infection has been associated with maternal anaemia, acute pyelonephritis, preterm labour, septicaemia and even possible death of the mother. Asymptomatic UTI can also cause intra-uterine growth restrictions, prematurity, and low birth weight of the fetus and sometimes even fetal mortality (9).

Recent WHO statistics show that 810 women die annually due to preventable causes related to pregnancy and childbirth (2). Majority (94%) of these maternal deaths occur in developing countries following poor access to healthcare services (10,11). Despite the standard diagnosis and management of UTI in-line with Ministry of Health (MOH) recommendations, current national statistics in Kenya show 488/100,000 maternal and 24/1000 neonatal mortality rates (12,13). This depicts a huge disparity in health service provision as compared to developed countries with minimal maternal mortality rate reported at 7/100,000 in 2017 (2,14). In Kenya, UTI was reported at a prevalence of 27% in the general population and it still remains of great public health importance (15,16). Gestational UTI contribute to substantive proportions of adverse pregnancy outcomes manifesting as different maternal and neonatal morbidity complications and deaths in Kenya (17,18). Regionally, North Eastern report the highest burden of maternal mortality rate (MMR) at 2,014/100,00 live births followed by Nyanza with 546/100,000, then, Eastern 400, Rift Valley 377, Coast 328, Western 319, Central 289, and Nairobi at 212/100,000 (2,19). These statistics vary in the different Counties within Kenya with Mandera having the highest reported MMR at 3,795, Wajir 1,687, Turkana 1,594, Marsabit 1,127, Isiolo 790, Siaya 691, Lamu 676, Migori 673, Garissa 646, Taita Taveta 603, Kisumu 597, Homa Bay 583 and Vihiga 531(12,20,21). A relative reduction in the burden of maternal mortality was realized following improved access to early screening and awareness, hence, in 2020, Kisumu County in western Kenya reported a maternal mortality rate of 495/100,000 still considerably high as compared to national statistics reported at 488/100,000 (22,23). The high burden of maternal mortality is directly attributed to haemorrhage (severe bleeding), obstetric labour, eclampsia, sepsis and raptured uterus; and indirectly attributed to illness aggravated by pregnancy including HIV/AIDs, anaemia, cardiovascular cause and malaria (24,25). Previous findings have reported 21.5% prevalence of UTI in pregnant women in Nairobi (16), 11.9% in children aged 2 months in rural settings (26,27). Previous findings (28) show a general association of UTI and pregnancy-related complications among women in Ethiopia. Untreated UTI infections pose a high risk of pyelonephritis, premature delivery and fetal mortality among pregnant women, these conditions are also associated with impaired renal function and end stage renal disease among paediatric patients (29,30). Although previous findings report pregnancy-related mortality (21,23,31,32), the prevalence rates of UTI associated maternal complications experienced annually are yet to be investigated. Early screening, improved hygiene and effective antibiotic treatment of gestational UTI hold a great potential to reduce maternal and neonatal morbidities and mortalities in Kisumu County. The need to understand the epidemiology and biology of UTI associated complications among pregnant women in Kisumu County calls for a robust data set to define UTI public health burden during gestation and the associated health complications. This study therefore, targeted Chulaimbo and Nyahera Sub-County hospitals as a model to establish UTI prevalence and specific clinical diagnosis and therapeutic management strategies among pregnant women seeking maternal child health (MCH) services from Chulaimbo and Nyahera Sub-County hospitals within Kisumu County.

## METHODS

### Study Area

This study was carried out in a hospital set-up within Kisumu County, Kenya. Kisumu County sits on a 2,085Km^2^ land, with a total population of 1,081,485, as at previous census done in 2018 hence a population density of 495 people/Km^2^. It is administratively divided into seven Sub-Counties including Kisumu Central (78,737 people), Kisumu East (69,988 people), Kisumu West (61,187 people), Nyando (65,751 people), Muhoroni (67,955 people), Seme (46,063 people) and Nyakach (62,024 people). Most residents in this county reside in the rural areas having 461,189 verses 429,354 people in the urban areas. Considering the environmental indicators, 52% of the total population access safe water while 47% have improved sanitation. Kisumu County reports a high fertility risk at 60% in women aged 20-29 and women younger than 20 years. Fertility risk inequalities are evident across all socio-demographic and socio-economic stratums. Health-seeking behaviour is affected by distance to the facility (33). In Kisumu County, 42% of women have distance issues to health service access, which are above the national rate at 20%. Kisumu County reports high (70%) ANC use as recommended by the WHO, however, only 27% receive all the components of ANC services including; blood pressure measurements, provision of a blood sample, provision of a urine sample, tetanus vaccination, Intermittent preventive treatment of malaria in pregnancy (IPTp), deworming treatment and iron-folic acid supplements (23). Maternal mortality rate (MMR) is 495/100,000 higher than the National MMR at 488/100,000, neonatal mortality rate (NMR) 39/1000 higher than the national NMR at 24/1000, child mortality rate (CMR) of 24/1000, infant mortality rate (IMR) of 54/1000 versus the National IMR at 31/100,000 and under-five mortality rate of 79/1000. Each Sub-County has a Sub-County level hospital with the capacity to offer ANC services, perform adequate microbiology diagnostic assays and run antibiotic susceptibility tests that reveal a high burden of multidrug resistance (MDR).

Data collection was targeted at Chulaimbo and Nyahera Sub-County hospitals in Kisumu East Sub-County since they contain the second largest population in the County. Furthermore, these hospitals serve a semi-urban population and have enhanced health services attributed to the presence of AMPATH program. These combinations of semi-urban location and having variety of other services qualified Chulaimbo and Nyahera Sub-County hospitals as the most appropriate data collection points for the current study.

### Study Design

This was a hospital-based study that adopted a prospective study design in which pregnant women’s hospital data extraction was done from health records over one year from February 2019 to February 2020.

### Study Population

The study population comprised of datasets from all women who attended MCH services from Chulaimbo and Nyahera Sub-County hospitals within Kisumu County from the month of February 2019 to February 2020 (See Supplementary Data I).

### Sample Population

The sample population included datasets from all pregnant women who attended MCH services from Chulaimbo and Nyahera Sub-County hospitals within Kisumu County from the month of February 2019 to February 2020.

### Inclusion criteria

Data from healthy pregnant women permanently residing in Kisumu County without any chronic infection, attending MCH services from Chulaimbo and Nyahera Sub-County hospitals and who presented with UTI infections at the hospital from February 2019 to February 2020.

### Exclusion criteria

Incomplete data sets from pregnant women and data from non-pregnant women were not eligible for the study.

### Sample size determination

Sample size was calculated using Fisher’s formula (34) at 50% UTI prevalence rate resulting N=365. As a precautionary measure, we considered an attrition rate of 10% (N=36.5) to give a final sample size of N=402. Therefore, in the current study, a total of 416 MCH datasets were collected over one year from February 2019 to February 2020.

### Sampling technique

Data from health records was extracted from Chulaimbo and Nyahera Sub-County hospitals records through purposive sampling.

### Quantitative data collection

Serialised electronic data capturing forms were used to collect socio-demographic and clinical data. Health records were interrogated to assess gestational UTI clinical diagnosis and therapeutic management.

### Qualitative data collection

Key informant interviews (KII) were conducted with 15 health care workers involved in direct care of pregnant women at Chulaimbo and Nyahera Sub-County hospitals. The interview guide focused on levels of UTI concern among health care workers, healthcare worker attention to pregnancy complications and health care worker directed UTI prevention and treatment strategies.

### Clinical Laboratory UTI diagnosis

As a way of confirming the specific bacteria associated with the UTI in the targeted population, a sub-set of 12 aseptically collected midstream urine samples were randomly selected from pregnant women attending ANC services from Chulaimbo and Nyahera Sub-County hospitals. These samples were microbiologically analysed and identified bacterial isolates subjected to antibiotic susceptibility tests (AST).

### Pilot testing

Prior to data collection, electronic data capturing forms were pretested on 10 MCH ANC data sets from Chulaimbo Sub-County hospital, which were not included in the final analyses.

Following piloting, these electronic data capturing forms were adjusted for clarity and consistency.

### Data Analysis

Data collected was recorded and safely saved in Excel spreadsheets. These were transferred to SPSS v21 database for analyses at the Department of Biomedical Sciences and Technology of Maseno University. Descriptive analysis was used to summarize the study population’s demographic characteristics. Chi-square tests were used for proportionality tests. A *P* ≤ 0.05 was considered statistically significant for all the analysis.

## RESULTS

The study population had a mean of two (2) ANC visits (±1.14; Min 1-Max 7); a mean mothers age of 23.92 years old (± 6; Min 10 – Max 43); a mean parity of 2 (±2; Min 0-Max 8); mean gravidae of 3 (± 2, Min 1-Max 10); mean gestational age of 24.5 weeks (±7.7 Min 3-max 38) and a mean haemoglobin level of 10.73(±1.8, Min 5 – Max 15). 56 % (233/416) attended the first ANC visit at varied gestational age during their pregnancy especially between 24 and 28 weeks **(Table 1)**. Only 1.4% (6/416) had a clinical history capturing UTI infection while a total prevalence of 57.9% (241/416) were UTI positive following their diagnosed routine ANC profile deep stick urinalysis test **(Table 2)**.

**Table 1:** Baseline demographic characteristics (N = 416)

| S/No | Variable | Mean | ±SD | Minimum | Maximum | Frequency | Prevalence (%) |
| --- | --- | --- | --- | --- | --- | --- | --- |
| 1 | No. of ANC visits | 2 | 1 | 1 | 7 |  |  |
| 2 | Mothers' Age (years) | 23.92 | 6 | 10 | 43 |  |  |
| 3 | Parity | 2 | 2 | 0 | 8 |  |  |
| 4 | Gravidity | 3 | 2 | 1 | 10 |  |  |
| 5 | Gestational age (weeks) | 24.52 | 7.7 | 3 | 38 |  |  |
| 6 | Haemoglobin level (g/dl) | 10.73 | 1.8 | 5 | 15 |  |  |
| 7 | First ANC visit |  |  |  |  | 233 | 56 |

**Table legend:** % Prevalence generated by proportionality tests using Ch-square. SD=Standard deviation; ANC=Ante-natal clinic.

**Table 2:** Clinical History and UTI status.

| S/No | Variable | Mean | ±SD | Minimum | Maximum | Frequency | Prevalence (%) |
| --- | --- | --- | --- | --- | --- | --- | --- |
| 1 | **Clinical History (N = 416)** |  |  |  |  |  |  |
|  | Clinical UTI diagnosed cases |  |  |  |  | 6 | 1.4 |
|  | Clinical UTI undiagnosed cases |  |  |  |  | 410 | 98.6 |
| 2 | **UTI infection status (deep stick urinalysis) (N= 416)** |  |  |  |  |  |  |
|  | Positive |  |  |  |  | 241 | 57.9 |
|  | Negative |  |  |  |  | 175 | 42.1 |

**Table legend:** % Prevalence generated by proportionality tests using Ch-square. SD=Standard deviation.

The proportion of pregnant women with asymptomatic UTI infections were57.9% (241/416), significantly high compared to those with negative UTI infection status 42.1% (175/416) (χ^2^, *p*<0.001). Clinical UTI history important for informed therapeutic management was performed for only 2.5% (6/241) cases who tested positive following deep stick urinalysis test. The remaining 97.5% (235/241) had no clinical history captured and no appropriate treatment administered. The datasets with clinical history captured revealed that a total of 1.4% (6/416) antibiotic therapy was mainly done by administration of first line broad-spectrum antibiotics.

Clinical laboratory diagnosis of specific aseptically collected midstream urine samples revealed the presence of Coagulase Negative *Staphylococcus*, Gram Positive Bacilli and *Streptococcus agalactae*. Antibiotic susceptibility tests showed likely resistance to commonly administered first line antibiotics including; ciprofloxacin, amoxicluvaic acid, ampicillin, tetracyclin, cetriaxone, sulfamethoxazole trimethoprim, ampicillin and gentamycin, but sensitivity to various antibiotics that are not available easily to such hospital set-ups such as Imepenem, erythromycin, chloramphenicol and clindamycin **(Table 3)**.

**Table 3:**
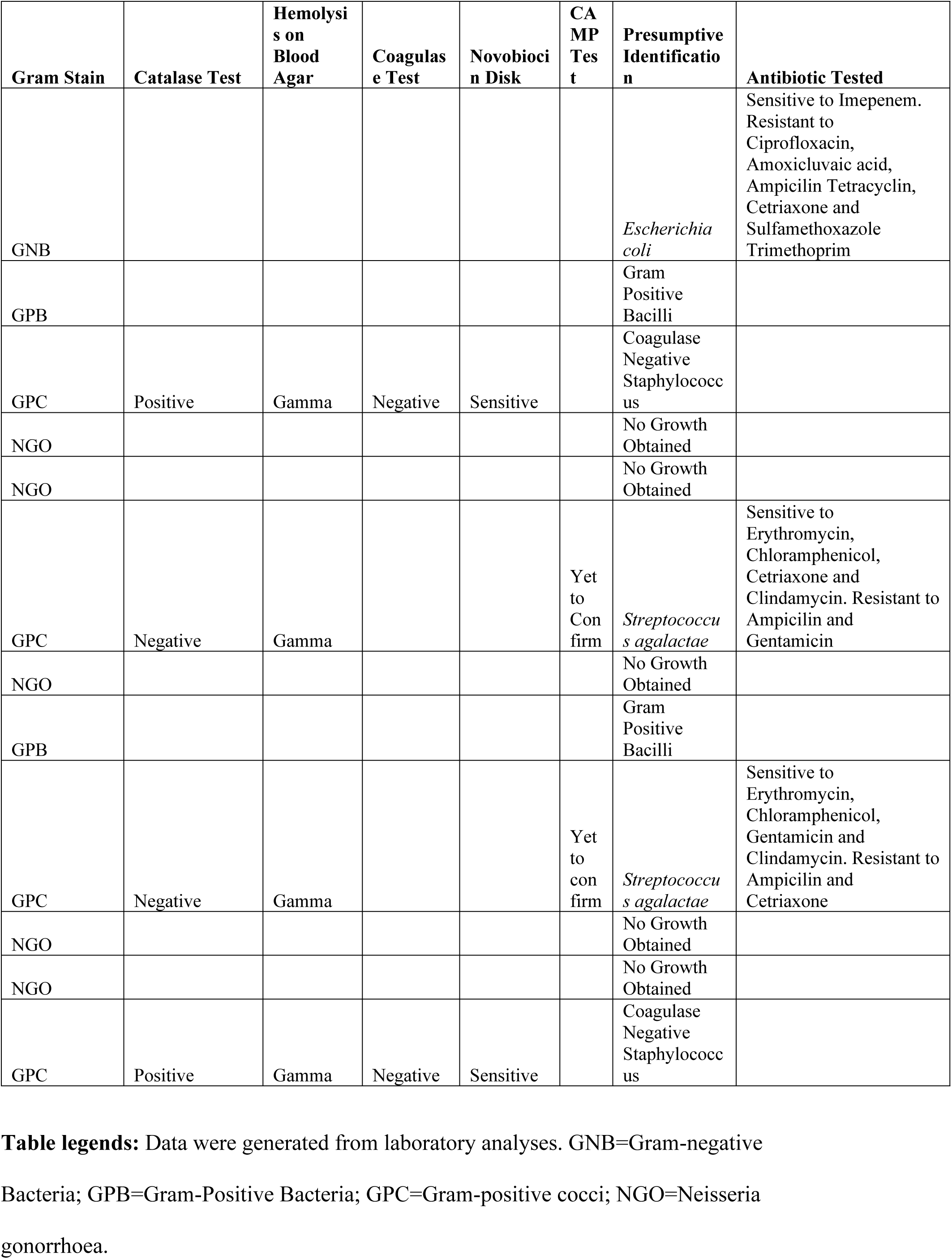
Laboratory Analysis of the UTI Specimens.

| Gram Stain | Catalase Test | Hemolysis on Blood Agar | Coagulase Test | Novobiocin Disk | CAMP Test | Presumptive Identification | Antibiotic Tested |
| --- | --- | --- | --- | --- | --- | --- | --- |
| GNB |  |  |  |  |  | *Escherichia coli* | Sensitive to Imepenem. Resistant to Ciprofloxacin, Amoxiclavate acid, Ampicillin Tetracyclin, Ceftriaxone and Sulfamethoxazole Trimethoprim |
| GPB |  |  |  |  |  | Gram Positive Bacilli |  |
| GPC | Positive | Gamma | Negative | Sensitive |  | Coagulase Negative Staphylococcus |  |
| NGO |  |  |  |  |  | No Growth Obtained |  |
| NGO |  |  |  |  |  | No Growth Obtained |  |
| GPC | Negative | Gamma |  |  | Yet to Confirm | *Streptococcus agalactiae* | Sensitive to Erythromycin, Chloramphenicol, Ceftriaxone and Clindamycin. Resistant to Ampicillin and Gentamicin |
| NGO |  |  |  |  |  | No Growth Obtained |  |
| GPB |  |  |  |  |  | Gram Positive Bacilli |  |
| GPC | Negative | Gamma |  |  | Yet to confirm | *Streptococcus agalactiae* | Sensitive to Erythromycin, Chloramphenicol, Gentamicin and Clindamycin. Resistant to Ampicillin and Ceftriaxone |
| NGO |  |  |  |  |  | No Growth Obtained |  |
| NGO |  |  |  |  |  | No Growth Obtained |  |
| GPC | Positive | Gamma | Negative | Sensitive |  | Coagulase Negative Staphylococcus |  |

**Table legends:** Data were generated from laboratory analyses. GNB=Gram-negative
Bacteria; GPB=Gram-Positive Bacteria; GPC=Gram-positive cocci; NGO=Neisseria
gonorrhoea.

Further qualitative results revealed that urinary tract infection during pregnancy was moderately of a concern among pregnant women seeking antenatal services at both facilities, and most (79% of the health workers) focused attention, on recurrent urinary tract infection and premature delivery as complications observed. Prevention strategies highlighted by most (63%) of the health workers to minimize the occurrence of urinary tract infection during pregnancy were education and counselling. In addition, majority (76%) of the health workers indicated that it is often, greatly emphasized, to pregnant women who test positive for a urinary tract infection that advice on personal hygiene and increasing water intake as the main ways of reducing UTI infections. Furthermore, most (71%) health care workers insisted on follow-up on the progress of those who test positive for a UTI at subsequent antenatal care visits. Finally, the specific management and treatment method for urinary tract infection highlighted by most (70% of health workers) was giving cefixime and nitrofurantoin based on physical assessment and personal judgment and assessment of severity. These antibiotics were considered in the absence of a drug susceptibility tests at the clinics.

## DISCUSSION

This study was set to determine the prevalence of gestational UTI, associated clinical diagnosis and the therapeutic management strategies. Socio-demographic, laboratory and clinical history data was extracted from 416 pregnant women’s MCH data sets in Chulaimbo and Nyahera Sub-County hospital facilities’ health records between February 2019 to February 2020 using pre-designed data collection forms.

Current research findings reveal asymptomatic gestational UTI prevalence of 57.9% (241/416) by routine empirical deep stick urinalysis test, which gives non-specific UTI associated aetiological bacterial agent findings. The current rate of gestational UTI prevalence in this region is much higher as compared to 27.6% reported in the general population among adults attending medical care in Kiambu level 5 hospital (15), and 21.5% among pregnant women attending ANC in Nairobi, Kenya (16). Other similar hospital-based studies in Sub-Saharan Africa have shown varying prevalence rates of gestational UTI infections; 3.7% Mbale, Uganda (35), 35% in Mbarara Uganda (36), 15.7 % in Ethiopia (28) and 20% and 35.5% in Tamale Central and Tamale Teaching hospitals in Ghana (37) and 18.8% in Sri Lanka, following urine culture and biochemical analysis during investigations. A systematic review of studies on the prevalence of gestational uro-pathogens and their antimicrobial resistance patterns in developing countries in Asia and Africa reported an overall rate of 13.5%. Gram-positive bacteria accounted for 15.9%, and Gram-negative bacteria accounted for the majority of infections (83.7%) with *Escherichia coli* being the most predominant uro-pathogen present in all the 26 studies included in the review. This review further reported resistance to antimicrobial drugs that are regularly used in developing countries with high resistance to ampicillin (67.2%). However, all the identified uro-pathogens showed relative sensitivity to ciprofloxacin (71.2%), nitrofurantoin (65%) and ceftriaxone (74.1%) (38). These previous observations are comparable to our findings, as their results shows that among pregnant women, the prevalence was at 55% in Benin City (39), and 56% in Onitsha (40), with an overall subtle variation in the rate of UTI in the different populations attributable to a variety of factors including parity, maternal age, gestational age, level of education and cultural practices.

The current study also shows significant omission of UTI status from the mothers’ clinical history following laboratory antenatal care (ANC) profiling. Only 2.5% (6/241) datasets had a clinical history capturing UTI infection out of the total prevalence of 57.9% (241/416) with positive UTI diagnosis by urine culture. A review of the diagnostic and therapeutic challenges in the management of gestational UTI revealed that specific detection and effective treatment remain a vital clinical problem in low-middle-income countries (LMICs) (30). In addition, previous works recommend repeated cultures every trimester for improved asymptomatic bacteriuria detection rate (41,42). Despite the Ministry of Health’s standard microbiological diagnostic procedures providing for bacterial culture and sensitivity tests, the LMICs have continued to carry out presumptive deep stick urinalysis for ANC profiling. These laboratory findings remain uncertain especially in regards to accurate identification of asymptomatic potentially pathogenic gestational bacteriuria. Inadequate relevant UTI history to clinicians hinders appropriate medication, hence, affected cases presenting with un-identified asymptomatic and un-characterised symptomatic UTI go un-noticed posing a future risk of gestational UTI associated complications. This is quite a worrying trend as the laboratory data in our settings demonstrated varied susceptibilities to various isolated pathogens. There is a general prescription of broad-spectrum antibiotics for UTI in this population and this might compromise future interventions due to suspected built up of resistant strains to the easily available broad-spectrum antibiotics.

Assessment of gestational UTI associated risk factors is highly advisable following the unmet challenges in the implementation of the ideal bacterial diagnostics for effective surveillance and proper clinical and therapeutic management (43).

Proper attention is missed out which may lead to recurrent UTI reinfections and ultimate life-threatening gestational UTI-associated complications. Previous review findings (44,45) reported no significant UTI association with primary pregnancy outcomes including pyelonephritis, preterm birth and secondary pregnancy outcomes including low birth weight, and Apgar score. On the contrary, some study findings report increasing positive association of gestational UTI with maternal anaemia, acute pyelonephritis, preterm labour, septicaemia and even possible death of the mother, intra-uterine growth restrictions, prematurity, and low birth weight of the fetus and fetal mortality (9). Gestational asymptomatic bacteriuria (ASB) has been associated with premature and low birth weight, a relationship that is not supported by other studies (46–48). Indeed, most of the healthcare workers in our set-ups mentioned some management strategies, which might be effective in reducing the prevalence of UTI in women. However, there were no clear follow-up strategies to ensure compliance and re-testing of the women who tested UTI positive. These mixed ideologies lead to controversial findings that require proper up-to-date implementation of the standard UTI diagnostic and management policy implementation for effective antibiotic therapy and ultimate healthy populations.

As previously stated above, misdiagnosis and inappropriate characterization of both asymptomatic and symptomatic UTI may lead to symptomatic treatment using broad-spectrum antibiotics posing a great risk for the development of multidrug resistant (MDR) bacterial strains (49,50). Previous findings reveal a future potential for the development of multidrug resistance (51) attributed to specific UTI bacterial aetiological agents (52,53) following poor clinical and therapeutic management. This may explain the increasing incidences of UTI associated complications in African populations where conclusive confirmatory tests as per the recommended Ministry of Health guidelines are not implemented due to limited resources.

## CONCLUSIONS

Current findings show a high prevalence of gestational UTI (57.9%). Only a handful of these cases get appropriate timely medication following clinical and therapeutic management. There is general lack of conclusive diagnostic test results in line with the Ministry of Health guidelines, which provide for bacterial culture and sensitivity tests. This leads to un-identification of UTI-specific aetiological bacterial agents prompting broad-spectrum antibiotic medications, a major risk for future antibiotic resistance, not conclusively addressed in the current study. There is paucity of information about common UTI associated aetiological bacterial agents in the target population. An investigation to identify specific UTI associated bacterial organisms will illuminate the possibility for development of alternative, more affordable point-of-care diagnostic platforms for enhanced UTI surveillance and specific antibiotic treatment (54,55). This will be vital in demystifying future forethought antibiotic resistance in populations at risk and ultimately a reduced burden of gestational UTIs and the associated poor pregnancy outcomes.

## Data Availability

Data will be available from the corresponding author upon request.

## LIST OF ABBREVIATIONS

AMPATH: Academic Model Providing Access to Healthcare
ANC: Antenatal Care
ASB: Asymptomatic Bacteriuria
AST: Antibiotic Susceptibility Test
CMR: Child Mortality Rate
GCRF: Global Challenge Research Fund
IMR: Infant Mortality Rate
IPTp: Intermittent Preventive Treatment of Malaria in Pregnancy
KII: Key Informant Interviews
LMIC: Low-and Middle-Income Countries
MCH: Maternal Child Health
MDR: Multidrug Resistance
MMR: Maternal Mortality Rate
MOH: Ministry of Health
NMR: Neonatal Mortality Rate
MUERC: Maseno University Ethical Review Committee
UTI: Urinary Tract Infections
WHO: World Health Organization

## DECLARATIONS

### ETHICAL APPROVAL AND CONSENT TO PARTICIPATE

The study protocol was approved by Maseno University Ethical Review Committee (MUERC) (Approval number MUERC/00959/21). An approval letter was also obtained from the Ministry of Health (MOH) local authorities for permission to access the hospital facility health records.

## CONSENT FOR PUBLICATION

Not applicable

## AVAILABILITY OF DATA AND MATERIALS

The datasets used during this study are available from the corresponding author on reasonable request.

## COMPETING INTERESTS

Not applicable

## FUNDING

De Montfort University QR-**Global Challenge Research Fund (GCRF) 2021**; Centre for Primary Healthcare Research. This funding body provided financial support for the collection, analysis and interpretation of data.

## AUTHORS’ CONTRIBUTIONS

ENT and SS conceptualised and designed the study. NTE, SS and EF, carried out data collection. NTE, SS, EF, TK, BO and CO analysed and interpreted data. All authors critically reviewed and approved the final manuscript for publication.

## ACKNOWLEDGEMENTS

We appreciate the Chulaimbo and Nyahera Suib-County hospital laboratory staff, clinical officers and the health records personnel for provision of the needed support and access to the data sets.

## HEALTH DATA PROTECTION

Health data was handled professionally and protected in line with the data protection Act No. 24 of 2019-Kenya Law. Data collected was securely stored in the Maseno University repository for future reference.

## Notes

### Competing Interest Statement

The authors have declared no competing interest.

### Funding Statement

None

### Author Declarations

Maseno University Ethics and Review Committee

## REFERENCES

1. Kisumu-County-Fact-Sheet-1-1.pdf [Internet]. [cited 2021 Dec 14]. Available from: https://www.kisumu.go.ke/wp-content/uploads/2018/11/Kisumu-County-Fact-Sheet-1-1.pdf

2. Maternal mortality rates and statistics [Internet]. UNICEF DATA. [cited 2021 Dec 13]. Available from: https://data.unicef.org/topic/maternal-health/maternal-mortality/

3. Koblinsky M, Chowdhury ME, Moran A, Ronsmans C. Maternal Morbidity and Disability and Their Consequences: Neglected Agenda in Maternal Health. J Health Popul Nutr. 2012 Jun;30(2):124–30.

4. Butler CC, Francis N, Thomas-Jones E, Llor C, Bongard E, Moore M, et al. Variations in presentation, management, and patient outcomes of urinary tract infection: a prospective four-country primary care observational cohort study. Br J Gen Pract. 2017 Dec;67(665): e830–41.

5. Rodriguez-Mañas L. Urinary tract infections in the elderly: a review of disease characteristics and current treatment options. Drugs Context. 2020 Jul 8;9: 2020–4–13.

6. Tonolini M, Ippolito S. Cross-sectional imaging of complicated urinary infections affecting the lower tract and male genital organs. Insights Imaging. 2016 Jun 7;7(5):689–711.

7. Vigil HR, Hickling DR. Urinary tract infection in the neurogenic bladder. Transl Androl Urol. 2016 Feb;5(1):72–87.

8. Nicolle L. Complicated urinary tract infection in adults. Can J Infect Dis Med Microbiol. 2005;16(6):349–60.

9. Gilbert NM, O’Brien VP, Hultgren S, Macones G, Lewis WG, Lewis AL. Urinary Tract Infection as a Preventable Cause of Pregnancy Complications: Opportunities, Challenges, and a Global Call to Action. Glob Adv Health Med. 2013 Sep;2(5):59–69.

10. Alkema L, Chou D, Hogan D, Zhang S, Moller A-B, Gemmill A, et al. National, regional, and global levels and trends in maternal mortality between 1990 and 2015 with scenario-based projections to 2030: a systematic analysis by the United Nations Maternal Mortality Estimation Inter-Agency Group. Lancet Lond Engl. 2016 Jan 30;387(10017):462–74.

11. Maternal mortality [Internet]. [cited 2021 Dec 10]. Available from: https://www.who.int/news-room/fact-sheets/detail/maternal-mortality

12. Kenya (KEN) - Demographics, Health & Infant Mortality [Internet]. UNICEF DATA. [cited 2021 Dec 14]. Available from: https://data.unicef.org/country/ken/

13. Maternal mortality ratio (modeled estimate, per 100,000 live births) - Kenya | Data [Internet]. [cited 2021 Dec 14]. Available from: https://data.worldbank.org/indicator/SH.STA.MMRT?locations=KE

14. Maternal mortality [Internet]. [cited 2021 Dec 13]. Available from: https://www.who.int/news-room/fact-sheets/detail/maternal-mortality

15. Wanja F, Ngugi C, Omwenga E, Maina J, Kiiru J. Urinary Tract Infection among Adults Seeking Medicare at Kiambu Level 5 Hospital, Kenya: Prevalence, Diversity, Antimicrobial Susceptibility Profiles and Possible Risk Factors. Adv Microbiol. 2021 Jul 29;11(8):360–83.

16. Ayoyi AO, Kikuvi G, Bii C, Kariuki S. Prevalence, aetiology and antibiotic sensitivity profile of asymptomatic bacteriuria isolates from pregnant women in selected antenatal clinic from Nairobi, Kenya. Pan Afr Med J. 2017 Jan 30; 26:41.

17. Wg M, Wp O, Tl H, J A. Contribution of urinary tract infection to the burden of febrile illnesses in young children in rural Kenya. Plos One. 2017 Mar 21;12(3): e0174199–e0174199.

18. Ayoyi AO, Kikuvi G, Bii C, Kariuki S. Prevalence, aetiology and antibiotic sensitivity profile of asymptomatic bacteriuria isolates from pregnant women in selected antenatal clinic from Nairobi, Kenya. Pan Afr Med J [Internet]. 2017 Jan 30 [cited 2020 Jul 26];26. Available from: https://www.ncbi.nlm.nih.gov/pmc/articles/PMC5398259/

19. Collaborators G 2015 MM. Global, regional, and national levels of maternal mortality, 1990–2015: a systematic analysis for the Global Burden of Disease Study 2015. Lancet Lond Engl. 2016 Oct 8;388(10053):1775.

20. Kenya mortality rate declines, CAS Health says – Ministry of Health [Internet]. [cited 2021 Dec 14]. Available from: https://www.health.go.ke/kenya-mortality-rate-declines-cas-health-says/

21. Finalpsareport_0.pdf [Internet]. [cited 2021 Dec 14]. Available from: https://www.unfpa.org/sites/default/files/admin-resource/Finalpsareport_0.pdf.

22. ete_of_rmncah_programme_report_online_2_0.pdf [Internet]. [cited 2021 Dec 14]. Available from: https://kenya.unfpa.org/sites/default/files/pub-pdf/ete_of_rmncah_programme_report_online_2_0.pdf.

23. fr308.pdf [Internet]. [cited 2021 Dec 14]. Available from: https://dhsprogram.com/pubs/pdf/fr308/fr308.pdf.

24. Filippi V, Chou D, Ronsmans C, Graham W, Say L. Levels and Causes of Maternal Mortality and Morbidity. In: Black RE, Laxminarayan R, Temmerman M, Walker N, editors. Reproductive, Maternal, Newborn, and Child Health: Disease Control Priorities, Third Edition (Volume 2) [Internet]. Washington (DC): The International Bank for Reconstruction and Development / The World Bank; 2016 [cited 2021 Dec 14]. Available from: http://www.ncbi.nlm.nih.gov/books/NBK361917/

25. Gülmezoglu AM, Lawrie TA, Hezelgrave N, Oladapo OT, Souza JP, Gielen M, et al. Interventions to Reduce Maternal and Newborn Morbidity and Mortality. In: Black RE, Laxminarayan R, Temmerman M, Walker N, editors. Reproductive, Maternal, Newborn, and Child Health: Disease Control Priorities, Third Edition (Volume 2) [Internet]. Washington (DC): The International Bank for Reconstruction and Development / The World Bank; 2016 [cited 2021 Dec 14]. Available from: http://www.ncbi.nlm.nih.gov/books/NBK361904/

26. Masika WG, O’Meara WP, Holland TL, Armstrong J. Contribution of urinary tract infection to the burden of febrile illnesses in young children in rural Kenya. Plos One. 2017 Mar 21;12(3): e0174199.

27. Sangeda RZ, Paul F, Mtweve DM. Prevalence of urinary tract infections and antibiogram of uropathogens isolated from children under five attending Bagamoyo District Hospital in Tanzania: A cross-sectional study [Internet]. F1000Research; 2021 [cited 2021 Dec 14]. Available from: https://f1000research.com/articles/10-449

28. Getaneh T, Negesse A, Dessie G, Desta M, Tigabu A. Prevalence of Urinary Tract Infection and Its Associated Factors among Pregnant Women in Ethiopia: A Systematic Review and Meta-Analysis. BioMed Res Int. 2021 Dec 1; 2021:6551526.

29. Tchente Nguefack C, Okalla Ebongue C, Nouwe Chokotheu C, Ebong Ewougo C, Nana Njamen T, Mboudou E. Clinical presentation, risk factors and pathogens involved in bacteriuria of pregnant women attending antenatal clinic of 3 hospitals in a developing country: a cross sectional analytic study. BMC Pregnancy Childbirth. 2019 Apr 29;19(1):143.

30. Matuszkiewicz-Rowińska J, Małyszko J, Wieliczko M. Urinary tract infections in pregnancy: old and new unresolved diagnostic and therapeutic problems. Arch Med Sci AMS. 2015 Mar 16;11(1):67–77.

31. ete_of_rmncah_programme_report_online_2_0.pdf [Internet]. [cited 2021 Dec 14]. Available from: https://kenya.unfpa.org/sites/default/files/pub-pdf/ete_of_rmncah_programme_report_online_2_0.pdf

32. Shrestha LB, Baral R, Poudel P, Khanal B. Clinical, etiological and antimicrobial susceptibility profile of pediatric urinary tract infections in a tertiary care hospital of Nepal. BMC Pediatr. 2019 Jan 29; 19:36.

33. Abubakar A, Van Baar A, Fischer R, Bomu G, Gona JK, Newton CR. Socio-Cultural Determinants of Health-Seeking Behaviour on the Kenyan Coast: A Qualitative Study. Plos One. 2013 Nov 18;8(11): e71998.

34. Self-designing clinical trials - Fisher - 1998 - Statistics in Medicine - Wiley Online Library [Internet]. [cited 2021 Mar 23]. Available from: https://onlinelibrary.wiley.com/doi/10.1002/(SICI)1097-0258(19980730)17:14%3C1551::AID-SIM868%3E3.0.CO;2-E

35. Nteziyaremye J, Iramiot SJ, Nekaka R, Musaba MW, Wandabwa J, Kisegerwa E, et al. Asymptomatic bacteriuria among pregnant women attending antenatal care at Mbale Hospital, Eastern Uganda. Plos One. 2020;15(3): e0230523.

36. Johnson B, Stephen BM, Joseph N, Asiphas O, Musa K, Taseera K. Prevalence and bacteriology of culture-positive urinary tract infection among pregnant women with suspected urinary tract infection at Mbarara regional referral hospital, South-Western Uganda. BMC Pregnancy Childbirth. 2021 Feb 23;21(1):159.

37. Karikari AB, Saba CKS, Yamik DY. Assessment of asymptomatic bacteriuria and sterile pyuria among antenatal attendants in hospitals in northern Ghana. BMC Pregnancy Childbirth. 2020 Apr 22; 20:239.

38. Belete MA, Saravanan M. A Systematic Review on Drug Resistant Urinary Tract Infection Among Pregnant Women in Developing Countries in Africa and Asia; 2005–2016. Infect Drug Resist. 2020 May 18; 13:1465–77.

39. Oladeinde BH, Omoregie R, Oladeinde OB. Asymptomatic Urinary Tract Infection among Pregnant Women Receiving Ante-Natal Care in a Traditional Birth Home in Benin City, Nigeria. Ethiop J Health Sci. 2015 Jan;25(1):3–8.

40. Prevalence of urinary tract infections in pregnant women in Onitsha, Nigeria. J Bacteriol Mycol Open Access [Internet]. 2018 Oct 12 [cited 2021 Dec 25]; Volume 6(Issue 5). Available from: https://medcraveonline.com/JBMOA/JBMOA-06-00219.pdf

41. McIsaac W, Carroll JC, Biringer A, Bernstein P, Lyons E, Low DE, et al. Screening for asymptomatic bacteriuria in pregnancy. J Obstet Gynaecol.

42. Tugrul S, Oral O, Kumru P, Köse D, Alkan A, Yildirim G. Evaluation and importance of asymptomatic bacteriuria in pregnancy. Clin Exp Obstet Gynecol. 2005;32(4):237–40.

43. Ngong IN, Fru-Cho J, Yung MA, Akoachere J-FKT. Prevalence, antimicrobial susceptibility pattern and associated risk factors for urinary tract infections in pregnant women attending ANC in some integrated health centers in the Buea Health District. BMC Pregnancy Childbirth. 2021 Oct 4; 21:673.

44. Schneeberger C, Geerlings SE, Middleton P, Crowther CA. Interventions for preventing recurrent urinary tract infection during pregnancy. Cochrane Database Syst Rev. 2015 Jul 26;2015(7):CD009279.

45. Lee AC, Mullany LC, Quaiyum M, Mitra DK, Labrique A, Christian P, et al. Effect of population-based antenatal screening and treatment of genitourinary tract infections on birth outcomes in Sylhet, Bangladesh (MIST): a cluster-randomised clinical trial. Lancet Glob Health. 2018 Dec 13;7(1): e148–59.

46. Chen Y-K, Chen S-F, Li H-C, Lin H-C. No increased risk of adverse pregnancy outcomes in women with urinary tract infections: a nationwide population-based study. Acta Obstet Gynecol Scand. 2010 Jul;89(7):882–8.

47. Schieve LA, Handler A, Hershow R, Persky V, Davis F. Urinary tract infection during pregnancy: its association with maternal morbidity and perinatal outcome. Am J Public Health. 1994 Mar;84(3):405–10.

48. Meis PJ, Michielutte R, Peters TJ, Wells HB, Sands RE, Coles EC, et al. Factors associated with preterm birth in Cardiff, Wales. II. Indicated and spontaneous preterm birth. Am J Obstet Gynecol. 1995 Aug;173(2):597–602.

49. Walker E, Lyman A, Gupta K, Mahoney MV, Snyder GM, Hirsch EB. Clinical Management of an Increasing Threat: Outpatient Urinary Tract Infections Due to Multidrug-Resistant Uropathogens. Clin Infect Dis. 2016 Oct 1;63(7):960–5.

50. Smith A, Anandan S, Veeraraghavan B, Thomas N. Colonization of the Preterm Neonatal Gut with Carbapenem-resistant Enterobacteriaceae and Its Association with Neonatal Sepsis and Maternal Gut Flora. J Glob Infect Dis. 2020 May 22;12(2):101–4.

51. Khawcharoenporn T, Vasoo S, Singh K. Urinary Tract Infections due to Multidrug-Resistant Enterobacteriaceae: Prevalence and Risk Factors in a Chicago Emergency Department. Emerg Med Int. 2013; 2013:258517.

52. Mutters NT, Mampel A, Kropidlowski R, Biehler K, Günther F, Bălu I, et al. Treating urinary tract infections due to MDR E. coli with Isothiocyanates – a phytotherapeutic alternative to antibiotics? Fitoterapia. 2018 Sep 1; 129:237–40.

53. Eshetie S, Unakal C, Gelaw A, Ayelign B, Endris M, Moges F. Multidrug resistant and carbapenemase producing Enterobacteriaceae among patients with urinary tract infection at referral Hospital, Northwest Ethiopia. Antimicrob Resist Infect Control. 2015 Apr 17; 4:12.

54. O’Brien VP, Hannan TJ, Nielsen HV, Hultgren SJ. Drug and Vaccine Development for the Treatment and Prevention of Urinary Tract Infections. Microbiol Spectr. 2016 Feb;4(1):10.1128/microbiolspec. UTI-0013–2012.

55. Davenport M, Mach KE, Dairiki Shortliffe LM, Banaei N, Wang T-H, Liao JC. New and developing diagnostic technologies for urinary tract infections. Nat Rev Urol. 2017 May;14(5):296–310.

